# Ketone-Based Therapies in Adults Heart Failure: A Systematic Review and Quantitative Analysis

**DOI:** 10.64898/2026.03.04.26347628

**Authors:** Aanchel Gupta, Yuliia Smereka, Wendimagegn Alemayehu, Robert Margaryan, Nariman Sepehrvand, Shubham Soni, Justin Ezekowitz

**Author notes:** Address for Correspondence: Justin Ezekowitz, MBBCh, MSc, 4-120 Katz Group Centre for Pharmacy and Health Research University of Alberta, Edmonton, AB, T6G 2E1. Relationships with Industry: No relationships to declare.

## Abstract

**Background:** Ketone bodies have shown potential to improve cardiac metabolism and function in patients with heart failure (HF).

**Objective:** To evaluate the effects of exogenous ketone-based interventions on cardiac function in patients with HF or related cardiometabolic risk factors.

**Methods:** We conducted a systematic review based on a search of MEDLINE, EMBASE, CINAHL, Cochrane Library, and Scopus from inception to January 2025. Eligible studies included randomized controlled trials evaluating exogenous ketones (oral ketones or ketone infusions) compared to placebo in adults with HF or patients with risk factors for HF including type 2 diabetes mellitus, hypertension, or coronary artery disease. Paired reviewers independently screened and identified hits at title-and-abstract and full-text levels to determine eligibility and extracted data from eligible studies. Random-effects meta-analysis was performed. Effects of interventions were summarized as mean differences (MD). Risk of bias was assessed using Cochrane RoB 2.0 tool. Certainty of evidence was evaluated using the GRADE (grading of recommendations assessment, development and evaluation) approach.

**Results:** Out of 565 unique records, 22 full-text articles were reviewed, and 8 studies met inclusion criteria. Exogenous ketone administration increased left ventricular ejection fraction (LVEF) (MD = 3.94, 95% CI 2.18-5.70, *p* = 0.001), cardiac output (CO) (MD = 1.11 L/min, 95% CI 0.55-1.67, *p* = 0.002), heart rate (4.85 bpm, 95% CI 2.24-7.46, *p* = 0.003), and stroke volume (SV) (MD = 10.21 mL, 95% CI 4.06-16.35, *p* = 0.005). Pulmonary capillary wedge pressure (PCWP) decreased (MD = -0.93 mmHg, 95% CI -1.44 to -0.43, *p* = 0.003), while mean arterial pressure showed no change (MD = -1.37 mmHg, 95% CI -3.53 to 0.79, *p* = 0.18).

**Conclusions:** Exogenous ketone-based therapies are associated with improvements in hemodynamic markers of cardiac function, including increases in LVEF, CO, and SV, along with a reduction in PCWP. These findings suggest that ketone supplementation may offer clinical benefits for patients with HF or vascular disease.

## Introduction

Heart failure (HF) is a global public health challenge, affecting an estimated 55 million individuals worldwide, corresponding to a prevalence of approximately 1-3% of the adult population.^1^ Between 2010 and 2019, the global prevalence of HF increased by nearly 30%, and remains a leading cause of hospitalizations and cardiovascular mortality.^2^ Guideline-directed medical therapy (GDMT) have proven mortality and morbidity benefits in HF with reduced ejection fraction (HFrEF), and more recently, in HF with preserved ejection fraction (HFpEF).^3^ However, despite optimal medical therapy, many patients remain symptomatic or at high risk of hospitalization. In addition, they have ongoing limitations such as issues related to cost, adherence, tolerability, and patient-specific contraindications may limit uptake.^4^ Thus, there remains an interest in non-traditional or adjunctive approaches.

As a result, there is growing interest in adjunctive therapies that may support cardiac function through alternative mechanisms. Nutraceuticals, defined as pharmacologically active compounds derived from food, are one such area of exploration.^5^ Among them, exogenous ketones have emerged as a possible option in HF management.^5,6^ Ketone bodies have recently emerged as a potential alternative therapeutic approach for the treatment of HF.^6–8^ Growing evidence has demonstrated that ketone bodies can both serve an additional metabolic substrate for the failing heart and have beneficial non-metabolic actions, such as vasodilation and reducing inflammation, all of which would be favorable for HF.^9,10^ Clinical studies using exogenous ketone supplementation, such as ketone esters, salts, or infusions, have demonstrated improvements in cardiac output (CO) and systemic hemodynamics, though evidence remains limited and heterogeneous.^11–13^

The objective of this systematic review is to synthesize the data from the studies that evaluated the effects of exogenous ketone supplementation in patients with HF or cardiometabolic risk factors. We aim to summarize the impact of ketone supplementation on cardiac function, hemodynamic parameters, and patient-important outcomes, and assess the overall quality of the current evidence.

## Methods

### Search Strategy

We searched MEDLINE, EMBASE, CINAHL, Cochrane Library, Scopus, and references of included studies dating from inception to January 2025. The search strategy combined key words and MeSH terms related to HF such as cardiovascular disease, ventricular dysfunction, and cardiac insufficiency with terms related to ketones including beta-hydroxybutyrate, and ketone esters. Specifically, we did not include ketogenic diets in our search terms. The protocol for the systematic review was registered with PROSPERO (CRD420261289091).

### Inclusion Criteria

Eligible studies were randomized controlled trials (RCTs), including crossover trials that compared exogenous ketones (including ketone salts, drinks or other oral supplements, infusions) with placebo. The population consisted of adult patients (>18 years) with HF or risk factors for HF, including type 2 diabetes mellitus, hypertension, or coronary artery disease (CAD). Studies were excluded if the population was healthy adults with no relevant comorbidities, if they evaluated ketogenic diets, raspberry ketones, or ginkgo extract, or if they lacked relevant cardiac function or hemodynamic outcomes. Finally, studies without a full text available, including relevant but ongoing trials, were excluded.

### Data Collection and Risk of Bias Assessment

Each title and abstract was screened by two authors and potentially relevant citations were retrieved and reviewed in full to determine eligibility. Extracted data included baseline characteristics such as age, sex, HF subtype if available, cardiovascular comorbidities, and acuity of presentation. The primary outcomes included measures of cardiac function including CO and left ventricular ejection fraction (LVEF). Additional outcomes included stroke volume (SV), heart rate (HR), pulmonary capillary wedge pressure (PCWP), and mean arterial pressure (MAP). We also included patient important outcomes such as all-cause mortality, hospitalizations, and emergency department visits. Other data extracted from the included studies related to the exogenous ketone intervention including dose, route of administration, duration of administration, and follow-up period. For studies with missing or incomplete outcome data, we contacted corresponding authors to request additional information.

Each included study was critically appraised by independent reviewers using the Cochrane Risk of Bias (RoB) 2.0 tool to assess the risk of bias in randomized trials.^14^ Reviewers rated trials as having a low risk of bias, probably low risk of bias, probably high risk of bias, or high risk of bias across the following domains: bias arising from the randomization process, bias owing to deviations from the intended intervention, bias from missing outcome data, bias in outcome measurement, bias in the selection of reported results, and bias from competing risks. Discrepancies between reviewers were resolved through discussion and, when necessary, by third party consensus.

### Statistical Analysis

Where sufficient data were available, quantitative synthesis was performed using random-effects meta-analysis to account for anticipated clinical and methodological heterogeneity across studies. Comparisons were conducted between ketone-based therapy and placebo groups for reported physiological outcomes, including CO, LVEF, CO, SV, HR, and PCWP. Continuous outcomes were pooled using mean differences (MDs) or standardized mean differences (SMDs), as appropriate, with corresponding 95% confidence intervals (CIs). Heterogeneity was assessed using chi-squared tests.

Given the small number of included studies, formal statistical assessment of publication bias was not performed, as this measure may be unreliable in analyses with limited study numbers. Patient-important clinical outcomes, including mortality and HF-related hospitalizations, were not reported in the included studies and therefore could not be analyzed quantitatively. Where meta-analysis was not feasible, results were summarized descriptively. Quantitative findings are presented using forest plots where applicable.

### Certainty of Evidence Assessment

We assessed the certainty of evidence using the GRADE approach. Certainty of evidence for each outcome was rated as high, moderate, low, or very low based on the domains of risk of bias, inconsistency, imprecision, indirectness, and publication bias (**Supplementary Appendix**). As fewer than ten studies were included in the pooled analysis, we did not assess publication bias statistically. However, to mitigate this potential source of bias, our search strategy included a comprehensive range of databases, clinical trial registries, and preprint servers to capture both published and unpublished studies. This approach reduces, but does not eliminate, the risk of publication bias, and was considered in the overall certainty of evidence assessment.

## Results

### Study Characteristics

The initial search resulted in 565 studies after duplicates were automatically removed. 23 full-text articles were reviewed, and 8 studies met inclusion criteria (**Figure 1**).

**Figure 1:**
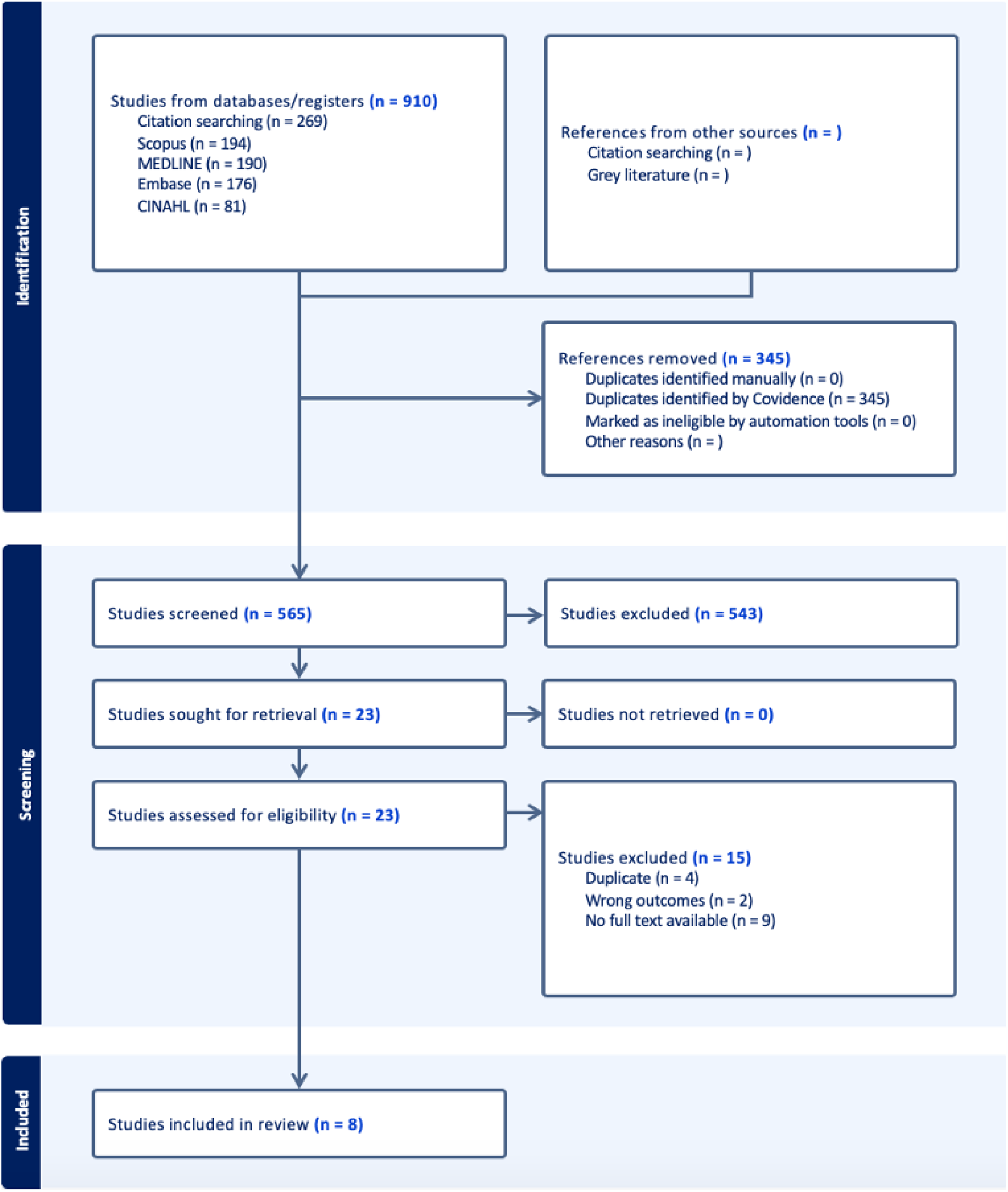
PRISMA (preferred reporting items for systematic reviews and meta-analyses) Flow Diagram of Selected Studies

Despite our effort, we were unable to access the data from Solis-Herrera for further analysis. Characteristics of the included studies are found in **Table 1**. All of the included studies were cross-over RCTs with washout periods between interventions. Study populations were small and between-study heterogeneity was present. All studies included patients with HF; however, some included those with additional comorbidities including diabetes and pulmonary hypertension. Interventions consisted of short-term ketone administration using varying formulations and doses, with follow-up ranging from acute measurements to 14 days.

**Table 1.** Characteristics of included studies.

| Author | Country | Participants | Setting | Route of therapy | Duration of therapy | HF subtype | Comorbidities |
| --- | --- | --- | --- | --- | --- | --- | --- |
| Gopalasingam 2024 | Denmark | 24 | Outpatient | PO | 4x daily 2 weeks | HFpEF | T2DM |
| Berg-Hansen 2024 | Denmark | 24 | Outpatient | PO | 4x daily 2 weeks | HFrEF |  |
| Berg-Hansen 2023 | Denmark | 12 | CCU (shock) | PO | 3 hours | HFrEF |  |
| Nielsen 2023 | Denmark | 20 | Outpatient | IV | 2 hours | HFpEF | Pulmonary HTN |
| Nielsen 2019 | Denmark | 16 | Outpatient | IV | 6 hours | HFrEF |  |
| Gopalasingam 2023 | Denmark | 12 | Outpatient | IV | 3 hours | HFrEF |  |
| Solis-Herrera 2022 | USA | 12 | Outpatient | IV | 6 hours | HFrEF | T2DM |
| Gulbrandsen 2024 | Denmark | 12 | Outpatient | PO | 6 hours | HFrEF |  |

Abbreviations: HF, heart failure; PO, prolonged oral; HFpEF, HF with preserved ejection fraction; T2DM, type 2 diabetes mellitus; HFrEF, HF with reduced ejection fraction; CCU, critical care unit; IV, intravenous; HTN, hypertension.

### Effects of the Intervention

Across the seven included studies, exogenous ketone supplementation was associated with significant improvements in several measures of cardiac function (**Table 2**, **Figure 2**).

**Figure 2.**
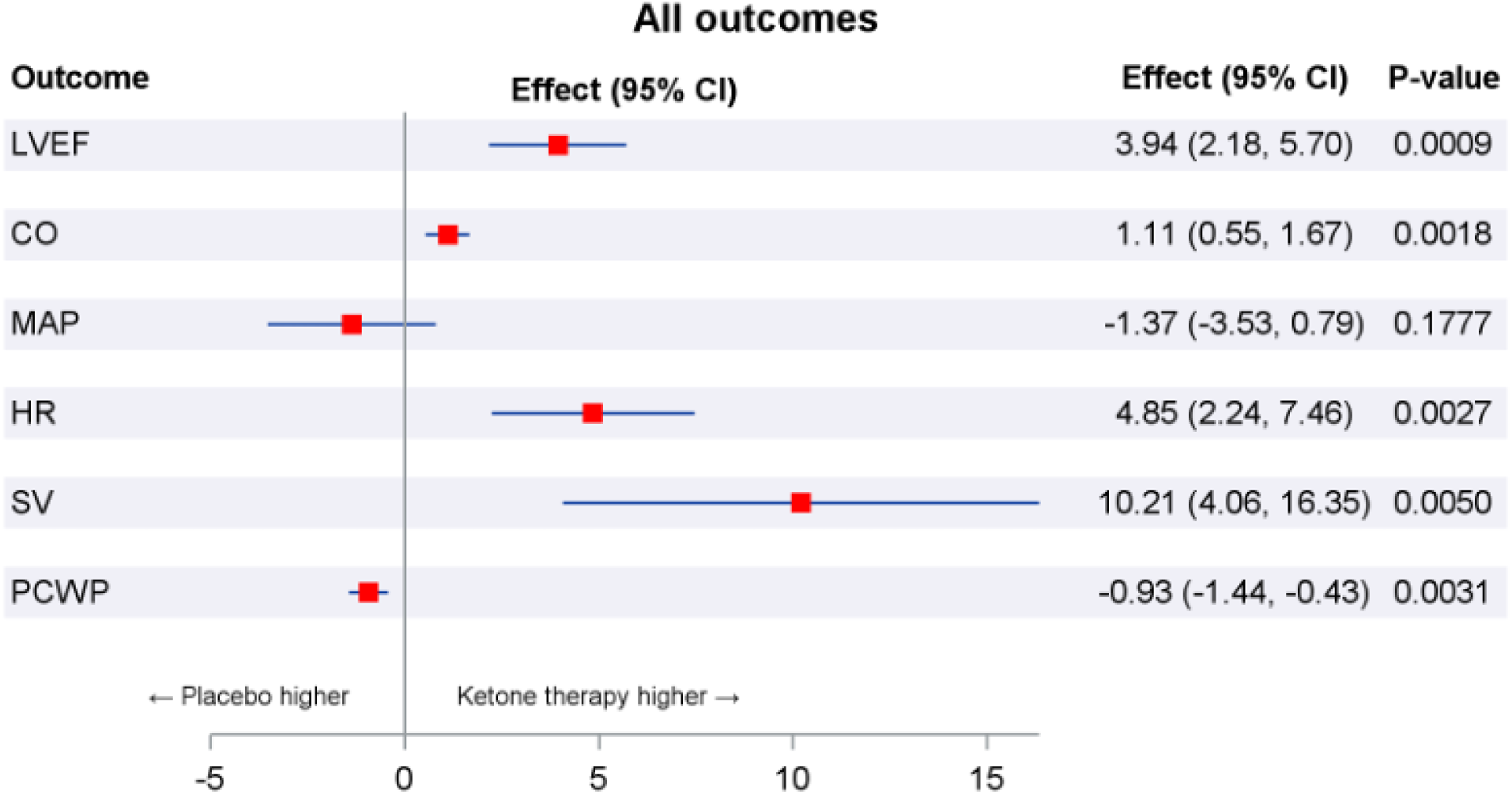
Forrest Plot of Random Effects Meta-analysis for Primary and Secondary Outcomes Abbreviations: CI, confidence interval; LVEF, left ventricular ejection fraction; CO, cardiac output; MAP, mean arterial pressure; HR, heart rate; SV, stroke volume; PCWP, pulmonary capillary wedge pressure.

**Table 2.**
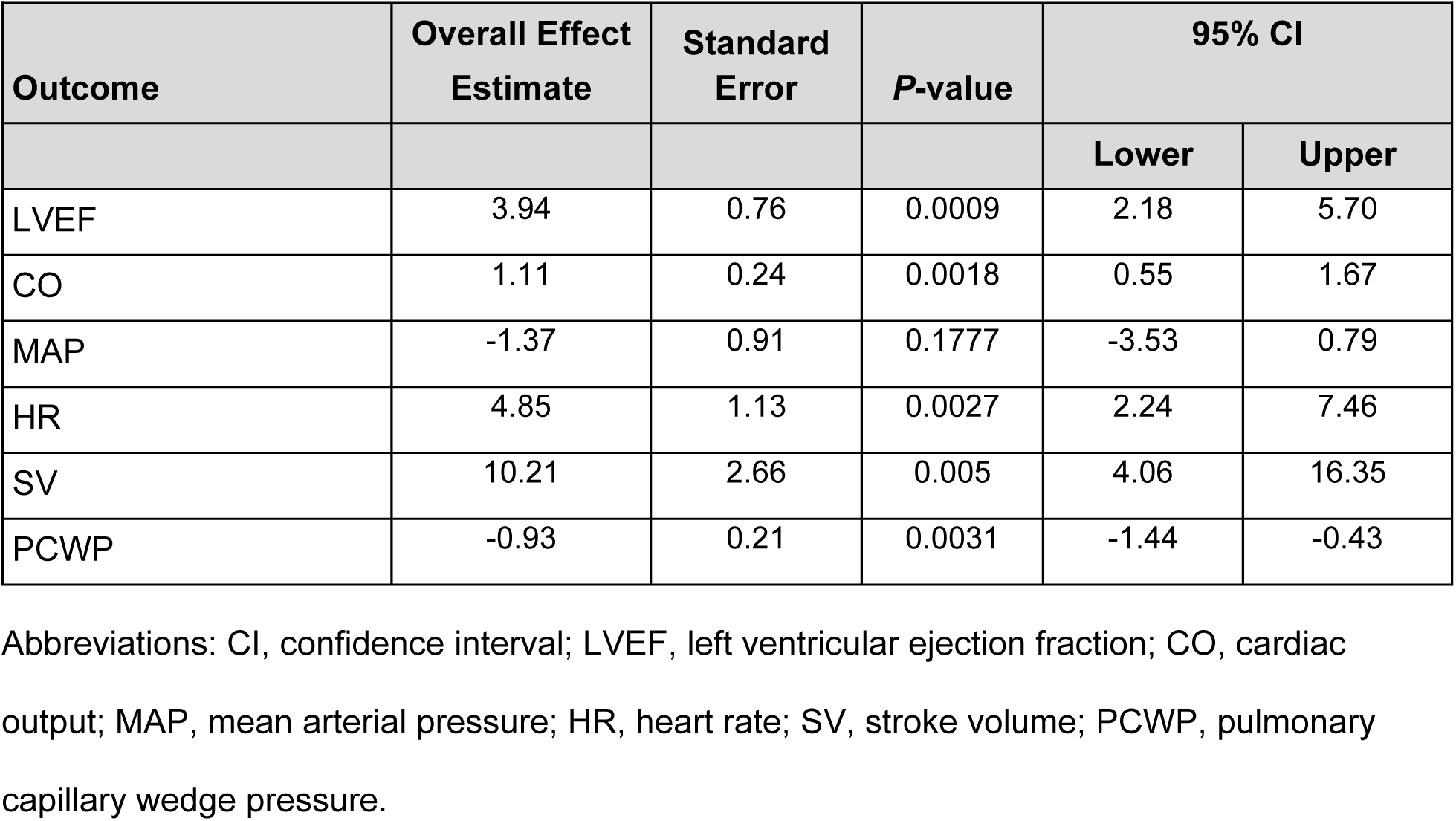
Summary Results of Random Effects Meta-analysis for Primary and Secondary Outcomes.

#### Left Ventricular Ejection Fraction

All of the studies reported LVEF with a maximum follow-up of 14 days. LVEF increased by a pooled mean difference of 3.94% with ketone administration (95% CI 2.18-5.70, *p* = 0.0009) (**Figure 3**). Substantial heterogeneity was present (I² = 77%), indicating considerable between-study variability. The certainty of evidence was moderate due to inconsistency pertaining to heterogeneity (**Table 3**).

**Figure 3.**
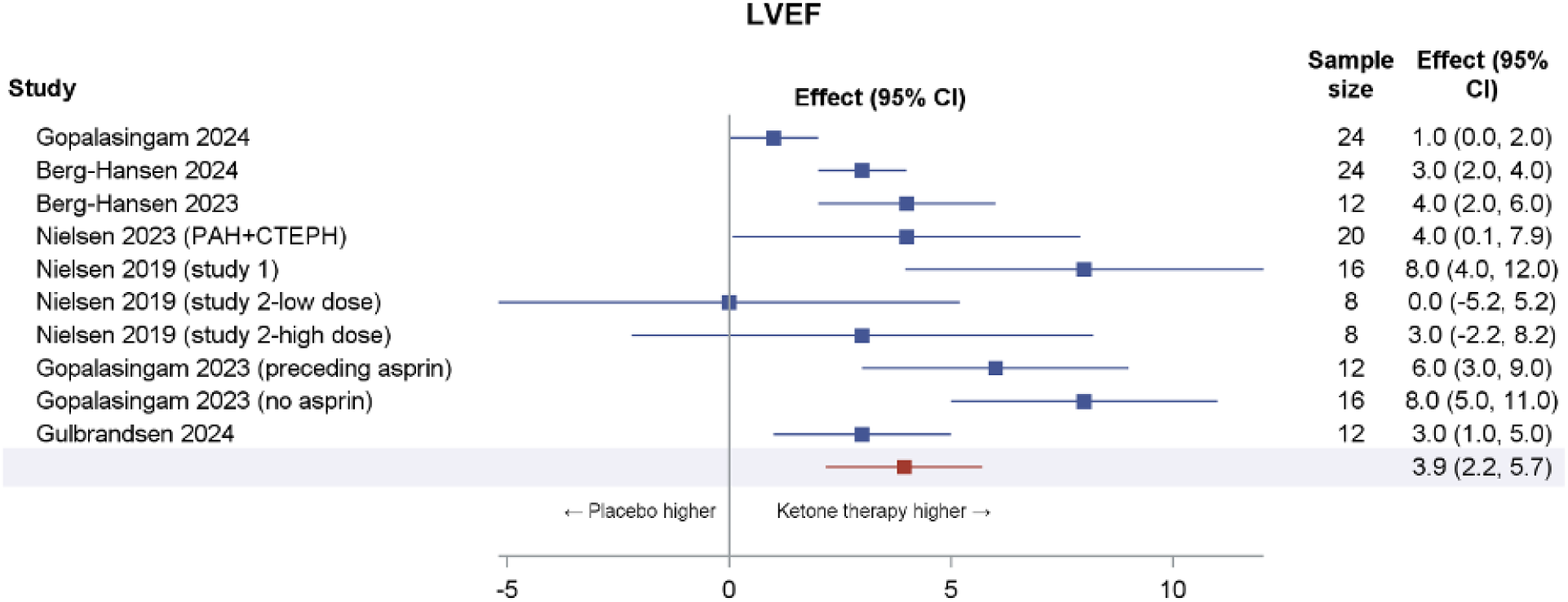
Forest Plot of LVEF Across Studies Abbreviations: LVEF, left ventricular ejection fraction; CI, confidence interval; PAH, pulmonary arterial hypertension; CTEPH, chronic thromboembolic pulmonary hypertension.

**Table 3.**
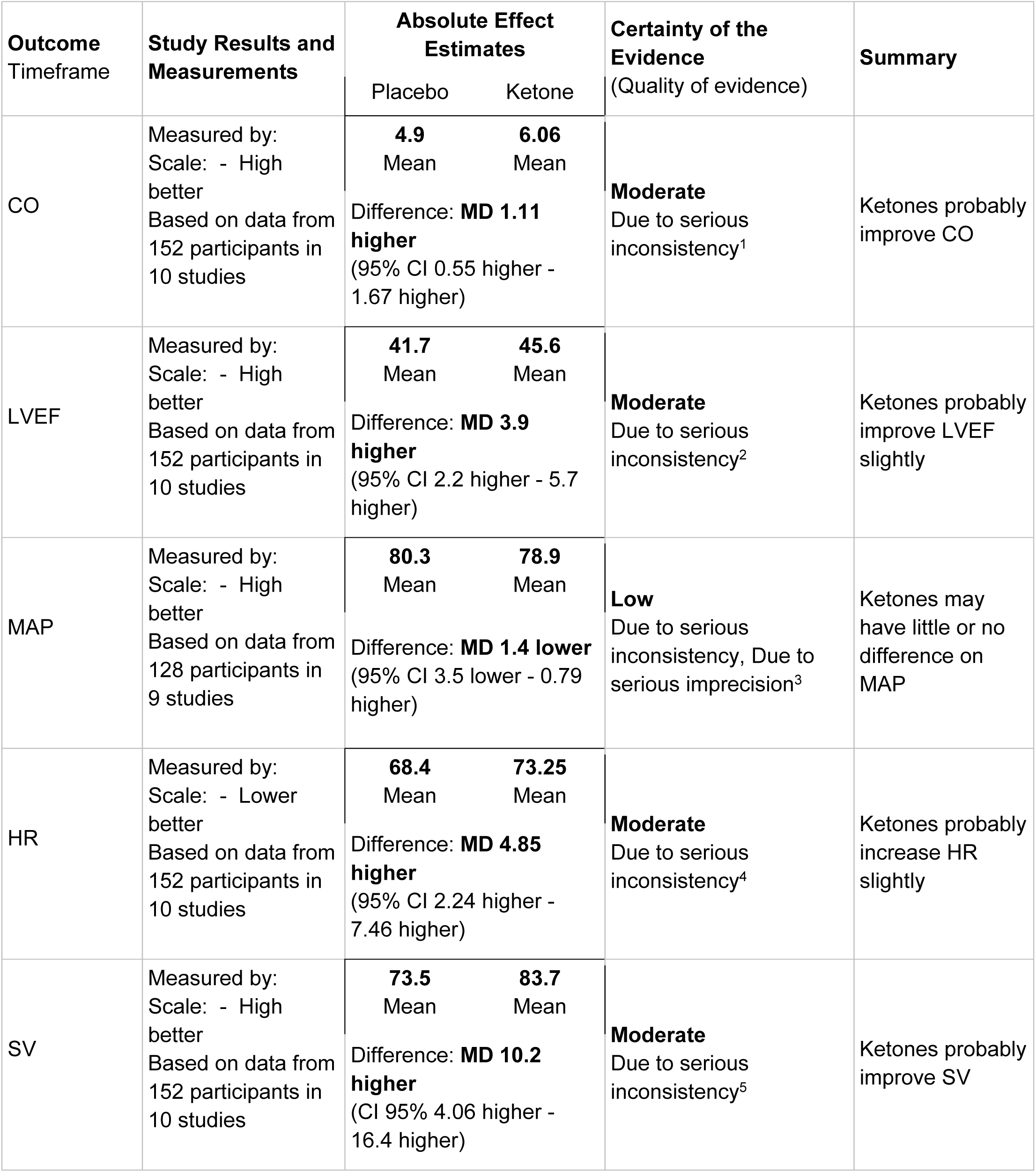
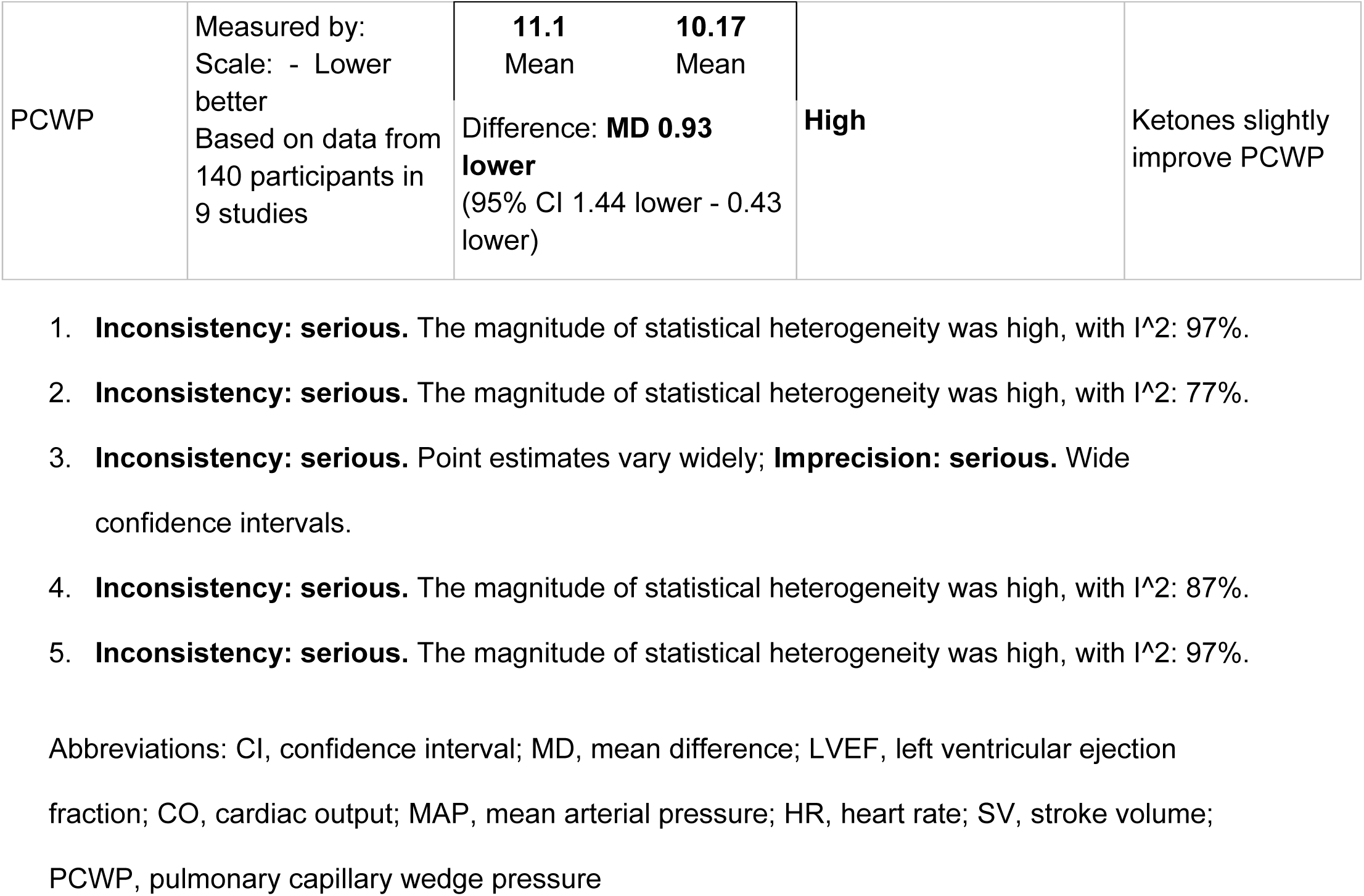
Summary of Findings Table.

#### Cardiac Output

Based on data from the seven studies and a total of 152 patients, CO was also significantly higher with ketone administration (MD 1.11 L/min, 95% CI 0.55-1.67, *p* = 0.0018) (**Figure 4**). Considerable heterogeneity was present (I² = 97%), indicating considerable between-study variability. The certainty of evidence was moderate due to serious inconsistency pertaining to heterogeneity (**Table 3**). The study by Nielson 2019 showed a significant difference in CO in the group that received the higher dose compared to the group that received a lower dose of ketones.

**Figure 4.**
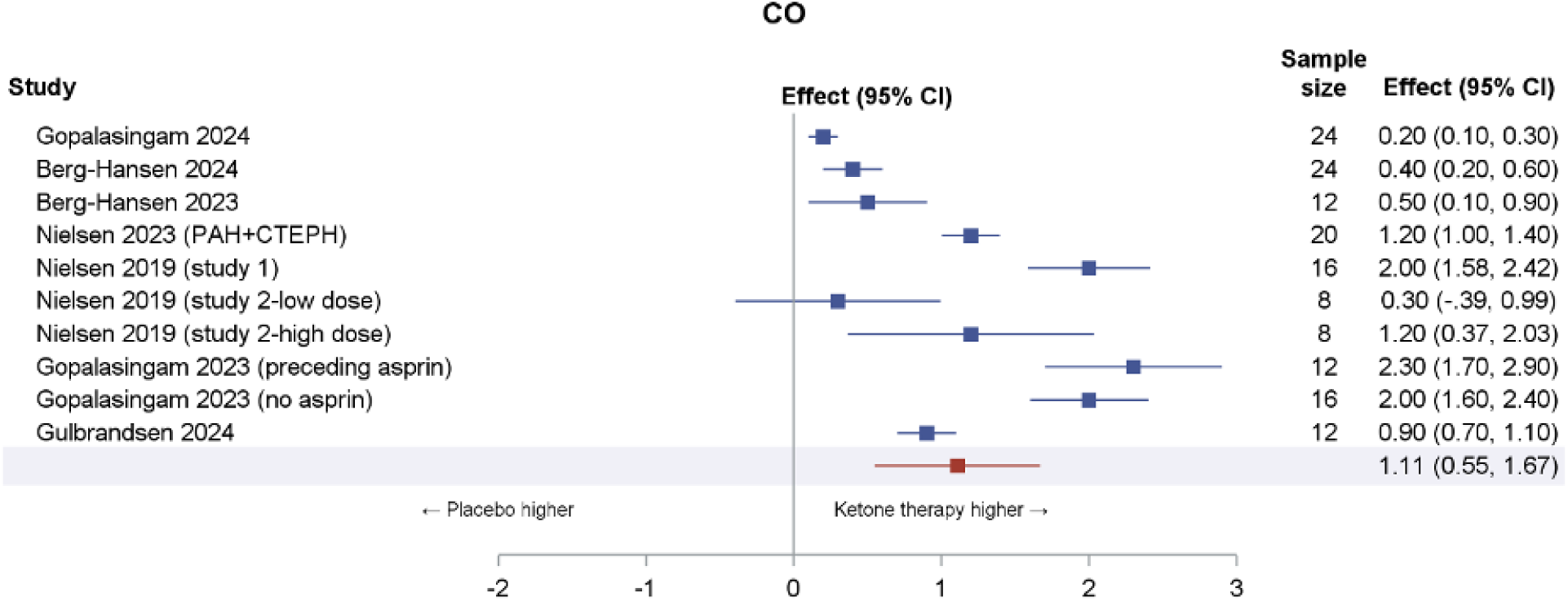
Forest Plot of CO Across Studies Abbreviations: CO, cardiac output; CI, confidence interval; PAH, pulmonary arterial hypertension; CTEPH, chronic thromboembolic pulmonary hypertension.

#### Stroke Volume

All of the studies reported SV with a maximum follow-up of 14 days. SV increased substantially with ketones (MD 10.21 mL, 95% CI 4.06-16.35, *p* = 0.005) (**Figure 5**); Considerable heterogeneity was present (I² = 94%), indicating considerable between-study variability; moderate certainty of evidence.

**Figure 5.**
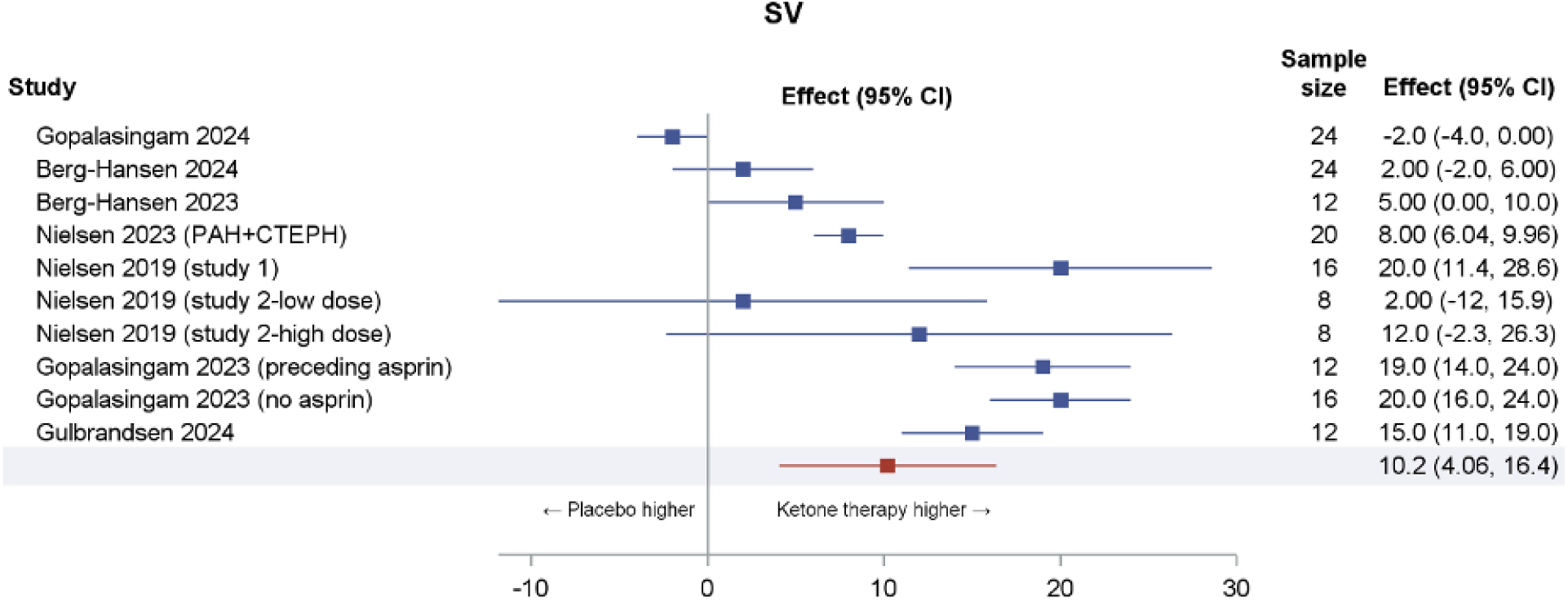
Forest Plot of SV Across Studies Abbreviations: SV, stroke volume; CI, confidence interval; PAH, pulmonary arterial hypertension; CTEPH, chronic thromboembolic pulmonary hypertension.

#### Heart Rate

Across all seven studies, HR rose by a mean of 4.85 beats per minute with ketone administration (95% CI 2.24-7.46, *p* = 0.0027) (**Figure 6**). Substantial heterogeneity was present (I² = 87%), indicating considerable between-study variability. Certainty of evidence was moderate due to imprecision related to heterogeneity.

**Figure 6.**
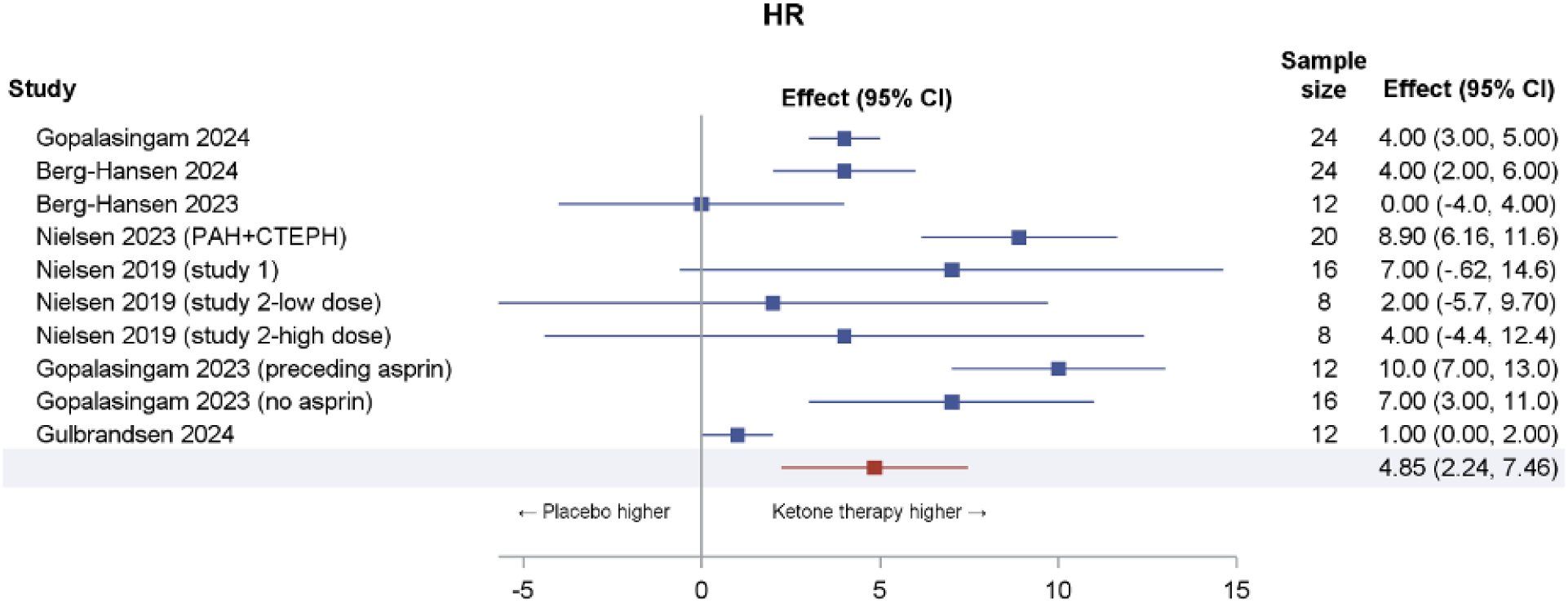
Forest Plot of HR Across Studies Abbreviations: HR, heart rate; CI, confidence interval; PAH, pulmonary arterial hypertension; CTEPH, chronic thromboembolic pulmonary hypertension.

#### Pulmonary Capillary Wedge Pressure

Based on data from six studies and a total of 140 patients, PCWP decreased significantly with ketone administration (MD -0.93 mmHg, 95% CI -1.44 to -0.43, *p* = 0.0031) (**Figure 7**). Between-study heterogeneity was low (I² = 0%), indicating consistent effects across studies. There is high certainty of evidence.

**Figure 7.**
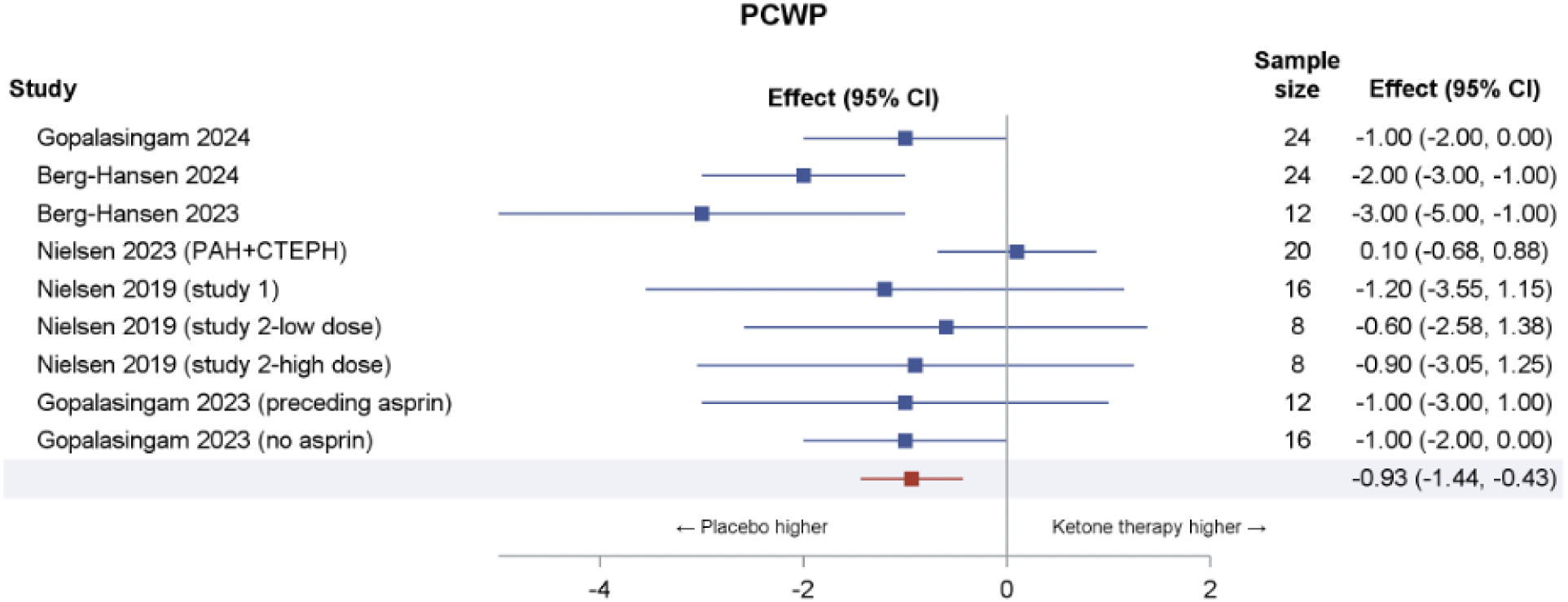
Forest Plot of PCWP Across Studies Abbreviations: PCWP, pulmonary capillary wedge pressure; CI, confidence interval; PAH, pulmonary arterial hypertension; CTEPH, chronic thromboembolic pulmonary hypertension.

#### Mean Arterial Pressure

Across the seven studies, no significant difference was observed for MAP with the ketone intervention (MD -1.37 mmHg, 95% CI -3.53 to 0.79, *p* = 0.18) (**Figure 8**). Moderate heterogeneity was observed (I² = 59%), suggesting some variability in effect estimates. Moderate certainty of evidence was observed due to imprecision related to statistical heterogeneity.

**Figure 8.**
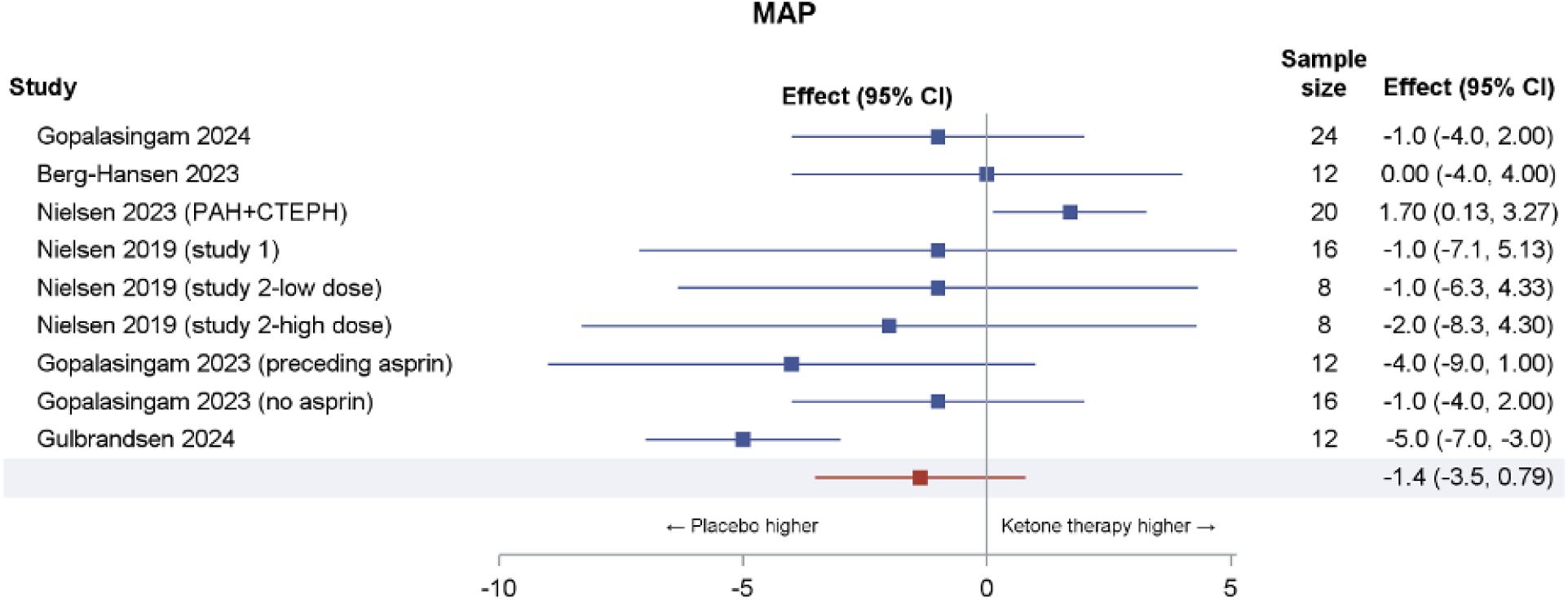
Forest Plot of MAP Across Studies Abbreviations: MAP, mean arterial pressure; CI, confidence interval; PAH, pulmonary arterial hypertension; CTEPH, chronic thromboembolic pulmonary hypertension.

### Risk of Bias of Included Studies

Risk of bias for each outcome can be found in supplementary materials. Overall, there was low risk of bias or some concerns across all outcomes. The studies with some concerns (Nielsen 2019, Gulbrandsen 2023) did not explicitly state whether allocation was concealed from those delivering the interventions, but the participants were blinded, and there did not appear to be significant difference in groups from period or sequence order. Additionally, in Guldbransen 2023, outcome assessors were aware of the intervention received by study participants.

The study by Solis-Herrera 2025 had some concerns of bias across all outcomes in the domains of bias arising from the randomization process, bias due to deviations from intended intervention, and bias in measurement of outcome. This study did not explicitly outline the randomization process, and the individuals delivering the intervention as well as measuring outcomes were aware of the assigned interventions.

## Discussion

This systematic review demonstrates that exogenous ketone therapy produces consistent short-term improvements in cardiac function and hemodynamics, including increases in LVEF, CO, and SV, and a reduction in PCWP. These changes were observed across different routes of administration, clinical settings, cardiometabolic comorbidities, and underlying HF phenotypes, such as HFpEF.

For most physiologic hemodynamic measurements, there are no universally accepted minimal clinically important differences (MCID), and when reported, they are usually disease or method specific. However, one study from 2011 measured clinically significant change in SV after a 6 minute walk test in patients with pulmonary hypertension, which was reported as ∼10ml.^15^ For LVEF, many HF studies and registries treated an absolute change of ≥5% in LVEF as clinically meaningful based on reproducibility thresholds of echocardiography, while larger absolute improvements (∼10%) correlated with measurable gains in functional status and quality of life.^16,17^ In contrast, MCIDs for CO and other secondary hemodynamic outcomes have not been consistently established, limiting interpretation of observed changes. The pooled LVEF improvement of ∼4%, with a confidence interval ranging from 2.1 to 5.7, in our analysis suggests that these changes may or may not be considered clinically relevant despite the improvements demonstrated in each individual study. Overall, these findings underscore the challenge of interpreting short-term physiologic changes in the absence of validated, outcome-linked clinical thresholds.

Ketone administration increased CO without affecting MAP, which distinguishes them from traditional inotropes. These results are similar to previous studies which show that ketone therapy can enhance cardiac contractility, induce vasodilation, and improve blood perfusion to the heart and other organs.^13,18,19^ Importantly, the observed hemodynamic profile differs from that of traditional inotropic agents, which often increase afterload or myocardial oxygen demand, suggesting a potentially more favorable energetic or vascular mechanism.^8,20^

The increase in HR observed across studies warrants cautious interpretation.

While short-term increases in HR may reflect a compensatory response accompanying improved CO, the long-term implications of sustained HR elevation in HF remain uncertain.^20^ Given the short duration of most included studies, it is not possible to determine whether these changes are transient, adaptive, or potentially maladaptive with prolonged exposure.

However, comparisons across studies are complicated by substantial heterogeneity in study populations, intervention protocols, and outcome assessment. The heterogeneity has also resulted in moderate certainty of evidence. Existing trials have enrolled patients with HFrEF, HFpEF, and cardiometabolic risk states, and have utilized varying ketone formulations, doses, and routes of administration, including both intravenous and oral preparations. These differences likely contribute to variability in observed effect sizes and limit the generalizability of pooled estimates.

Although a growing number of clinical trials are evaluating ketone supplementation in HF, consensus regarding their long-term role in disease management has not yet been established. Ongoing trials across the United States, Canada, Denmark, and The Czech Republic are investigating oral and IV ketone therapy in various HF settings. One study (NCT06937320) in Denmark aims to investigate longer term oral ketones in patients with HFpEF over 8 weeks. A number of trials including the KETO-AHF trial are investigating ketones in the acute HF or cardiogenic shock population (NCT06653725^21^, NCT04442555, NCT04698005). The present findings support the biological plausibility of these investigations and highlight the need for longer-duration trials powered to assess patient-important outcomes, safety, and durability of effect.

The available evidence suggests that exogenous ketones may have short-term hemodynamic or symptomatic effects, but the clinical relevance of these findings remains uncertain. These results support continued evaluation of ketone supplementation across other heart-failure phenotypes, including HFpEF, where metabolic impairment is also implicated and therapeutic options remain limited. Orally administered formulations may offer greater feasibility, scalability, and acceptability for outpatient management compared with intravenous delivery. Additionally, studies are needed to assess the longer-term effects of exogenous ketones, as existing trials have evaluated only acute or very short-term use. Well-designed studies with sufficient follow-up will be important to clarify the safety profile, durability of effect, and potential clinical impact of exogenous ketone therapy in HF.

### Strengths and Limitations

This review has several strengths. We conducted a comprehensive and methodologically rigorous search, with duplicate screening for eligibility, risk-of-bias assessment, and data extraction. We formally applied the GRADE framework to evaluate the certainty of evidence.

The review had potential limitations. The included studies were limited by the number of patients (maximum of 24 patients per study cohort), which limited the total sample size. The review was also limited by the duration of follow-up as most studies reported a one-time intervention of ketones. Only two studies examined ketone therapy daily for 14 days, limiting our analysis of therapy to the acute and sub-acute term. While all of the included studies examined patients with HF, they were also somewhat heterogeneous in terms of patient populations with additional comorbidities; Nielsen 2023 examined patients with HF and pulmonary hypertension, and Gopalasingam 2024 examined those with HF and type 2 diabetes. All but one study examined patients with stable HF in the outpatient setting, Berg-Hansen 2023 investigated patients with cardiogenic shock and HF in a cardiac intensive care setting.

None of the included studies evaluated patient-important outcomes such as mortality, hospitalizations, or quality of life. Instead, the available evidence focused primarily on short-term surrogate measures including CO, ejection fraction, and hemodynamic parameters. While these markers provide insights into the potential role of exogenous ketones, they do not establish whether supplementation translates into meaningful clinical benefit for patients living with HF, the mechanisms for which have been previously suggested.^22^ This gap highlights the need for adequately powered, longer-term trials that are designed to assess outcomes valued by patients and clinicians, such as symptom burden, functional capacity, and health-related quality of life.^23^

## Conclusion

Overall, the available evidence suggests that exogenous ketone therapy exerts favorable short-term hemodynamic effects across diverse HF populations. However, the absence of long-term data and clinically meaningful outcomes limits the ability to determine whether these improvements are sufficient to influence disease trajectory or patient well-being. High-quality randomized trials addressing these gaps are warranted.

## Data Availability

All data produced in the present study are available upon reasonable request to the authors.

