## Supplementary Appendix for "Ketone-Based Therapies in Adults Heart Failure: A Systematic Review and Quantitative Analysis"

**Figure 1.** Risk of Bias for Left Ventricular Ejection Fraction

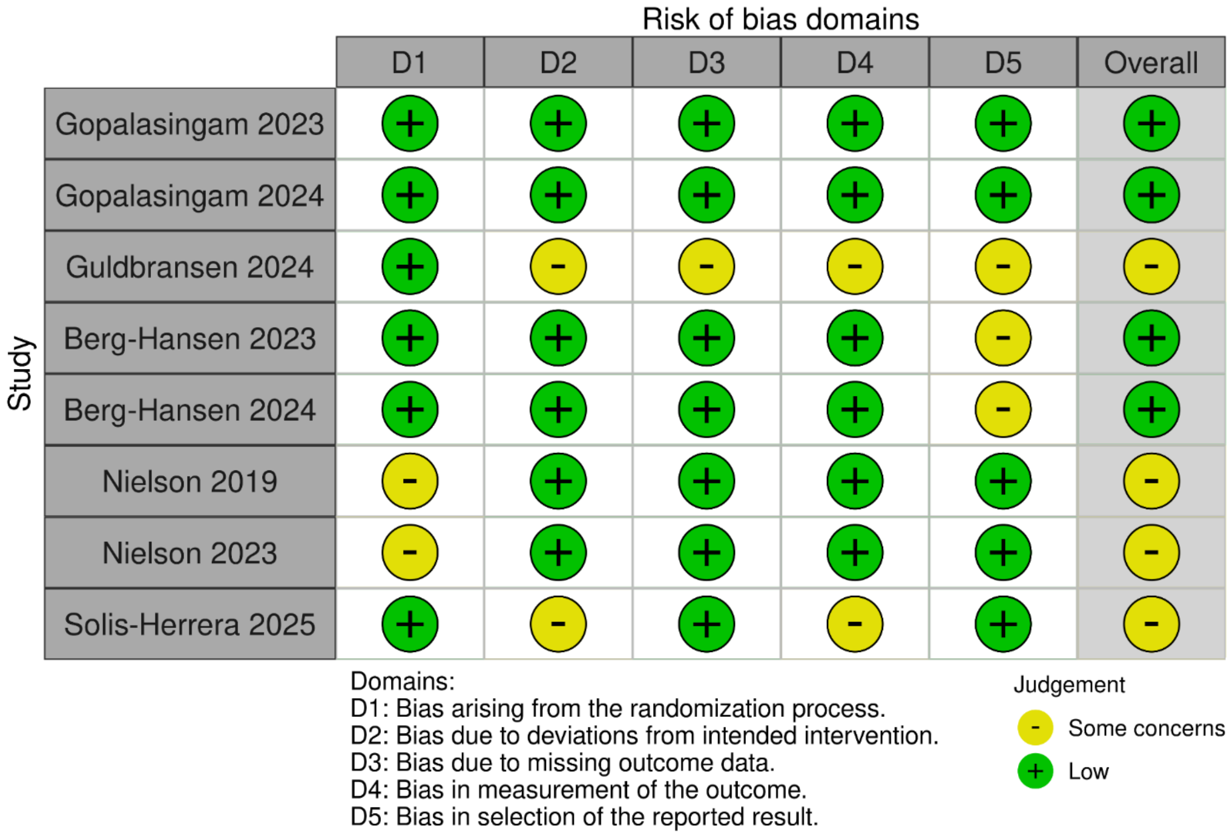

**Figure 2**. Risk of Bias for Cardiac Output

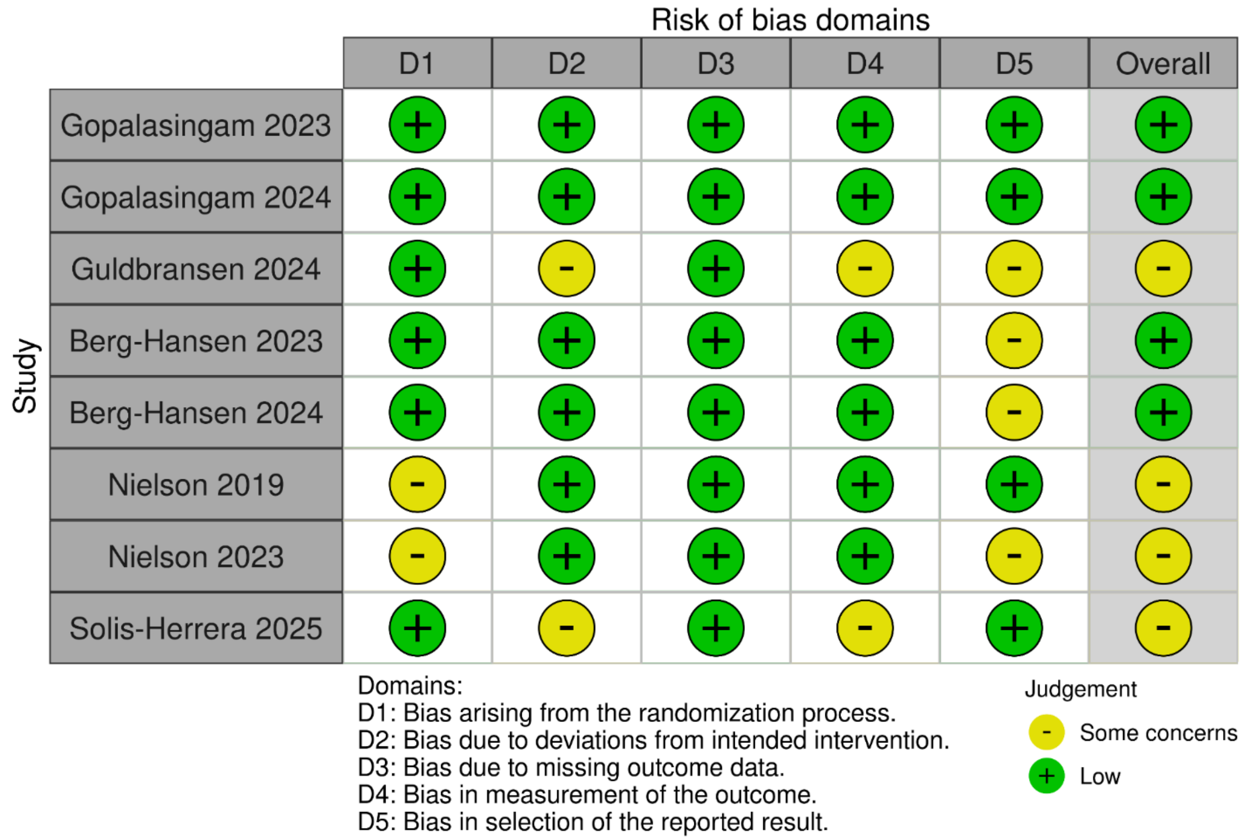

**Figure 3**. Risk of Bias for Mean Arterial Pressure

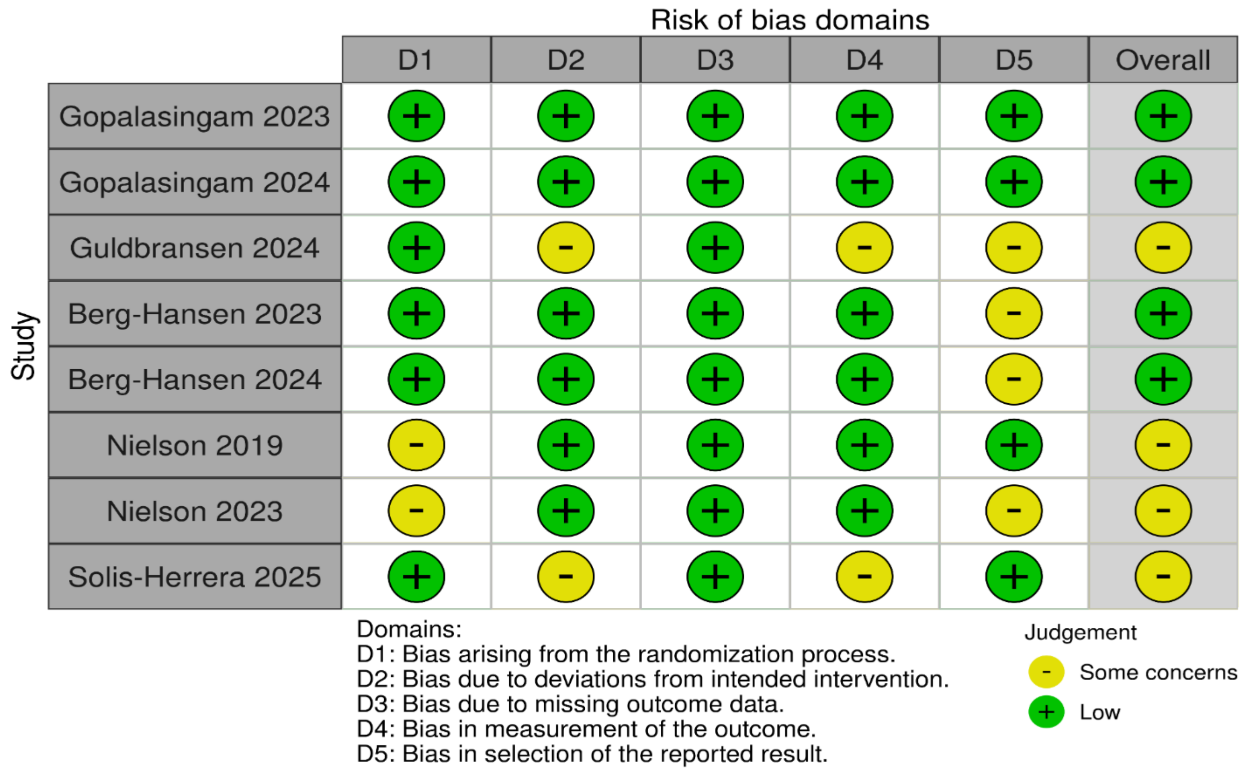

**Figure 4.** Risk of Bias for Stroke Volume

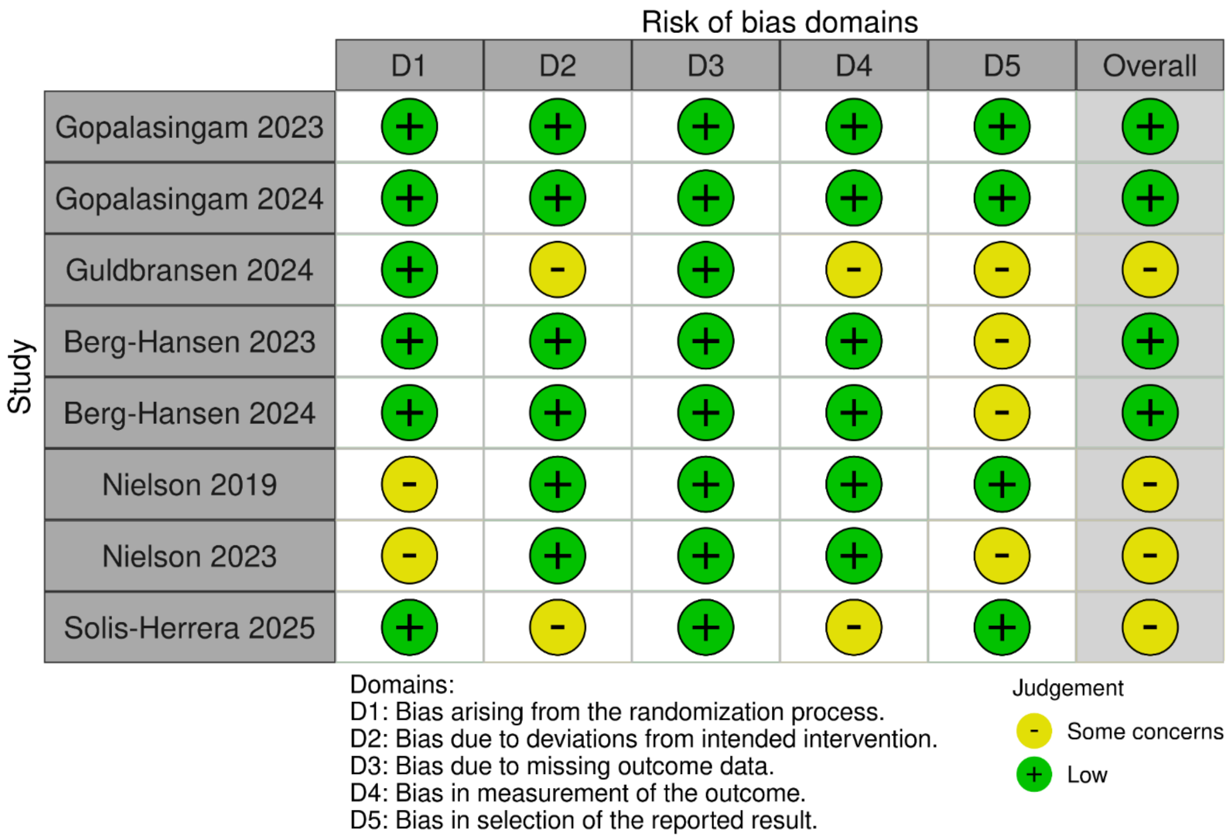

**Figure 5**. Risk of Bias for Heart Rate

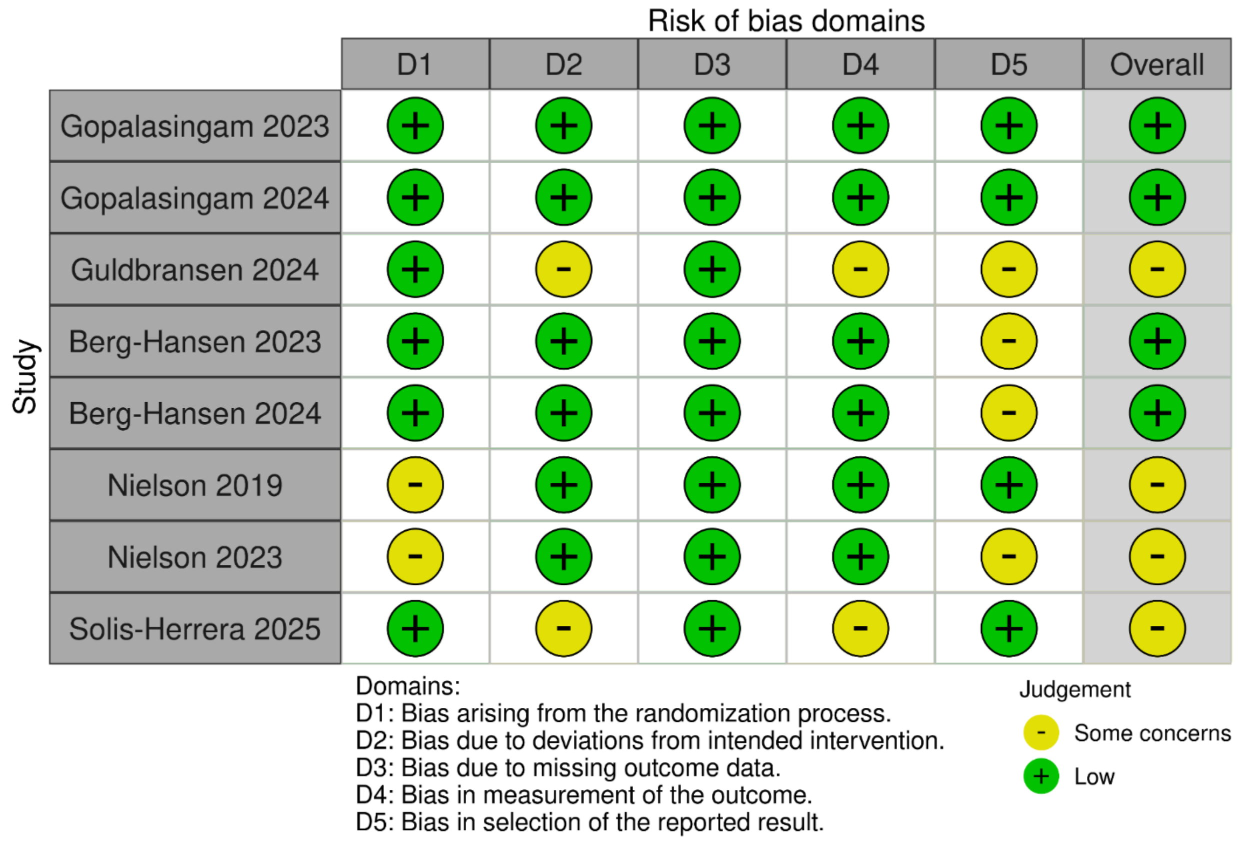

**Figure 6.** Risk of Bias for Pulmonary Capillary Wedge Pressure

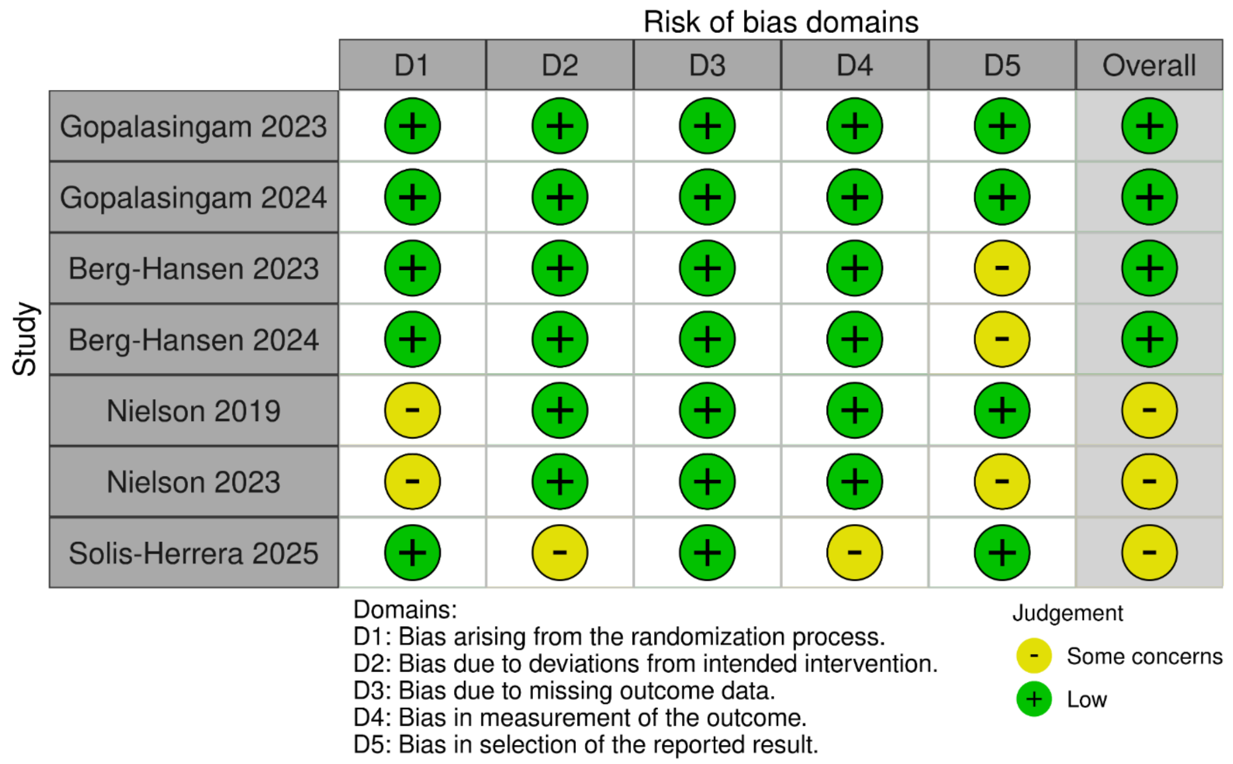
